# Ownership of mainstream media devices and digital access tools predict health insurance coverage status among Kenyans: A cross-sectional study of the Kenya Demographic and Health Survey (KDHS) 2022 dataset

**DOI:** 10.1101/2025.06.18.25329901

**Authors:** Elham Aldousari, Maha Alhajeri, Dennis Kithinji

## Abstract

**Background:** Low health insurance coverage, poverty, and reduced donor funding impede access to healthcare in Kenya. The Social Health Authority (SHA) unveiled in Kenya in October 2024 can provide the healthcare financing solutions needed through targeted interventions to improve health insurance coverage. This study sought to identify modifiable factors that influence health insurance coverage status in Kenya to provide the basis for targeted interventions by SHA.

**Methods:** The Household Recode, Individual Recode, and Men’s Recode datasets from the Kenya Demographic and Health Survey conducted between February and July 2022 were combined to form the dataset. Proportions of individuals with and without health insurance as well as those with the various potential determinants were calculated. The associations between the potential determinants and health insurance coverage status were estimated using bivariate analysis through Pearson’s Chi-square test. Multivariable logistic regression facilitated the identification of determinants of health insurance coverage in Kenya.

**Results:** Data of 14232 participants aged 15-54 years whose health insurance coverage status were indicated were analyzed. The participants were mainly female (66%), in good health (79%), literate (75%), relatively poor (56%), connected to electricity (55%), and radio listeners (61%). The rate of health insurance coverage was 34%, with 93% of them covered by NHIF. Out of the 16 potential predictors (chi-square range = 21-2694, p < 0.0001) included in the logistic regression model, only six were significant predictors of health insurance coverage: education level, wealth index, ownership of a solar panel, and television, mobile phone, and computer ownership (14-47% difference in odds).

**Conclusion:** Health insurance coverage remains low in Kenya. Education levels, economic status, and media factors are significant determinants of health insurance coverage. SHA can leverage the identified determinants to strengthen interventions that worked for NHIF and address the challenges that impede health insurance uptake among Kenyans in the informal sector.

## Introduction

Health insurance uptake in Kenya has been low, with only about 20% having health insurance coverage based on the 2014 Kenya Demographic and Health Survey (KDHS) (Kazungu & Barasa, 2017). Majority of the regular contributors to Social Health Insurance Fund (SHIF) are formal employees since employers are mandated to submit their contributions to the Social Health Authority (SHA) (Indimuli et al., 2023). Unfortunately, only about 23% of Kenyan workers are in the formal sector (Murunga et al., 2021), hence the government intervenes to convince Kenyans in the informal sector to regularly contribute to social health insurance (Barasa et al., 2018). Most Kenyans in the informal sector rely on out-of-pocket payments to access healthcare services (Salari et al., 2019). Yet, majority cannot afford quality healthcare services since 80% of Kenyans either live below the national poverty line or are income poor (Diwakar & Shepherd, 2018). Therefore, access to healthcare services is limited for most Kenyans.

The need for a functional and equitable social health insurance in Kenya increased in 2025 due to the budget cuts for USAID and the World Health Organization, which have been supporting several health programs in Kenya. The US government paused aid that led to discontinuation of PEPFAR in January 2025, and five countries that fund 90% of HIV care globally announced aid reductions ranging from 8%-70% (Ten Brink et al., 2025). Kenya seeks to leverage the SHA that replaced National Health Insurance Fund (NHIF) to finance healthcare through the premium-based Social Health Insurance Fund (SHIF) and tax-funded Primary Healthcare Fund (PHF) and the Emergency, Chronic, and Critical Illness Fund (ECCIF) (Nungo et al., 2024). The challenges that bedeviled NHIF are likely to persist in SHA if the underlying factors are not identified and addressed.

In the 2014 KDHS, older age, male gender, marriage, belonging to a small household, belonging to a rich household, having a chronic disease, and exposure to media were associated with health insurance coverage (Kazungu & Barasa, 2017). Another KDHS was conducted in 2022, hence analyzing the data to identify more specific and modifiable factors behind health insurance coverage is necessary. It can inform the design of interventions to increase individual contributions into the recently launched SHIF.

KDHS 2022 reported that only about 20% of Kenyans aged 15-54 years had more than secondary education, although over 90% were literate based on either having post-secondary education or ability to read a sentence presented to them (Kenya National Bureau of Statistics et al., 2023). Radio and television were the commonest means of media exposure, with more than 60% of Kenyans either listening to radio or watching television. More than half of the Kenyans use the internet on a daily basis (Kenya National Bureau of Statistics et al., 2023). Therefore, education level, listening to radio, watching television, and browsing the internet may be influencing their insurance coverage.

According to KDHS, only 26% of Kenyans had health insurance in 2022, majority being urban dwellers (40%) compared to rural residents (19%). Households in urban areas had higher rates of health insurance coverage (46% in Nairobi City County) compared to households in rural areas (5% in Tana River County) (Kenya National Bureau of Statistics et al., 2023). Cash payment was the main approach to incurring health expenditure (KSh. 13621 per year per household), followed by NHIF payments (KSh. 9330 per year per household), with urban dwellers making approximately twice as much cash payments as rural residents for health services (Kenya National Bureau of Statistics et al., 2023). This study aimed to identify modifiable predictors of health insurance coverage among Kenyans by analyzing the KDHS 2022 data.

## Methods

### Setting

This study was conducted in Kenya, a developing country that transitioned from NHIF to SHA to advance toward Universal Health Coverage in October 2024. The KDHS 2022, which is the 7^th^ and the most recent KDHS as at May 2025 when this study was conducted, collected data from 17^th^ February 2022 to 31^st^ July 2022 (Kenya National Bureau of Statistics et al., 2023).

### Study design

A cross-sectional study design was conducted using the Kenya Demographic and Health Survey (KDHS) 2022 data to estimate the level of insurance coverage among Kenyans and identify modifiable predictors of health insurance coverage in Kenya. The items that STROBE Statement indicates should be included in cross-sectional studies’ reports were optimally encompassed in this study.

### Participants

Data from individuals aged 15-54 years in households across Kenya, whose health insurance coverage data and data on its potential determinants were collected in KDHS 2022, were included in the study. Two-stage stratified sampling design with stratification for rural and urban residence was used in KDHS 2022. The sample in KDHS 2022 included 1691 clusters across Kenya, 666 in urban areas and 1025 in rural areas selected using Equal Probability Selection Method (EPSEM). Twenty-five households were selected per cluster; a total of 42022 households were included since some clusters had fewer than 25 households.

### Variables

The dependent variable was the health insurance status of the individuals. The independent variables were education attainment and indicators of access to media (having electricity, radio, television, telephone, mobile phone, computer, and internet access).

Indicators of access to media could be critical determinants of health insurance status since the Ministry of Health in Kenya and other stakeholders use mass media and digital media to communicate health insurance messages (Kenya National Bureau of Statistics et al., 2023).

### Data source – retrieval of KDHS 2022 data

KDHS 2022 data were obtained from Kenya National Data Archive (KeNADA) for analysis (Kenya National Bureau of Statistics (KNBS), 2023). Data on health expenditure and insurance cover were collected from 21996 households. The Household Recode (household characteristics), Individual Recode (women aged 15-49 years), and Men’s Recode (men aged 15-54 years) datasets were combined to form the dataset that was analyzed in this study. KDHS created the recodes from the data collected through computer-assisted personal interviews in preselected households.

### Bias

Sampling in the KDHS 2022 was done without replacement to avoid duplication, which would have reduced the representativeness of the sample and introduced bias in the estimation of population parameters.

### Ethical considerations

This study analyzed an anonymized Public Use Dataset from the Kenya Demographic and Health Survey (KDHS) collected and maintained using strict ethical guidelines by the Kenya National Bureau of Statistics (KNBS) in collaboration with international partners. A request to access the KDHS 2022 data for a research study was made to the KNBS and granted. All the access conditions by KNBS were observed. Authors did not have access to information that could identify individual participants during or after data collection.

### Data analysis

Frequencies of health insurance coverage and its potential determinants were obtained in counts and percentages. Bivariate analysis was conducted using Pearson’s Chi-square to estimate the association between the potential determinants and status of health insurance coverage. All significant modifiable factors (P < 0.05) were included in a multivariable logistic regression model since the outcome was binary (having or lacking a health insurance cover) to identify determinants of the status of health insurance. Pearson’s R correlation coefficient (r ≥ 0.8) were assessed to identify multicollinearity before fitting the model.

## Results

### Characteristics of the participants

Out of the 14232 respondents aged 15-54 years old whose status of health insurance coverage were indicated in the KDHS 2022 dataset, 37.0% were covered by health insurance (table 1). Their mean age was 32.9 years (standard deviation, SD = 9.1).

**Table 1.**
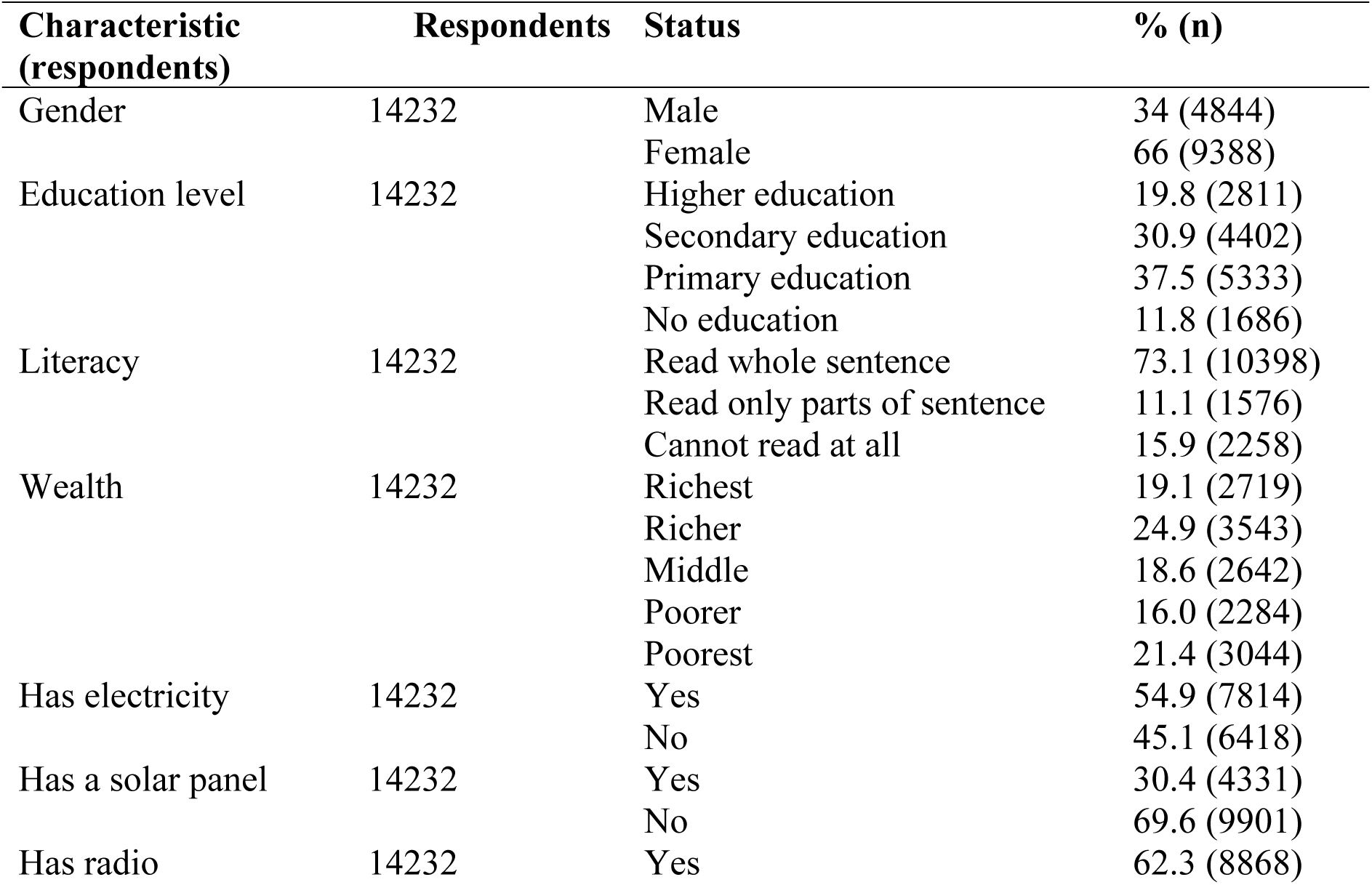
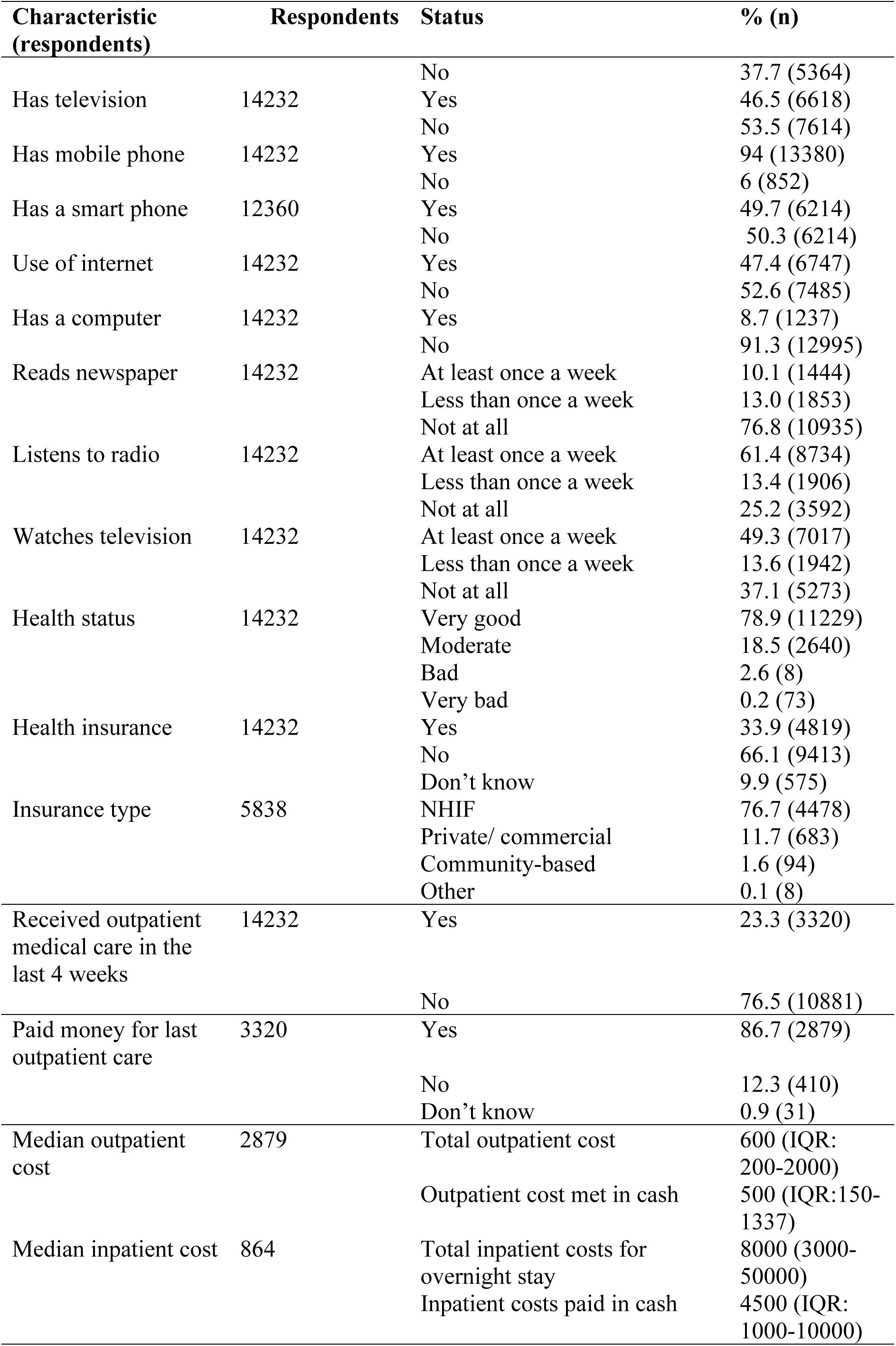
Demographic, socioeconomic, and health characteristics of the respondents covered by health insurance.

The total outpatient costs varied significantly from as low as zero (n = 58) to as high as KSh.530000 (n = 1). Only 3.1% (n = 84) of them exceeded KSh. 10000. Cash and NHIF were the main approaches to meeting outpatient healthcare costs as 91% of the 2698 respondents paid some cash (KSh.1 - 130000) for healthcare costs; 56 of the cash payments being KSh. 10000 or more. NHIF paid bills for 5.6% (n = 148) respondents (range: KSh.100 - 400000); only 18 were KSh. 10000 or more. Eight respondents indicated that the amount was met in kind (range KSh. 500 - 20000). Fifty-two respondents had their amounts covered by private insurance (range KSh. 20 - 100000), with only 10 getting payments exceeding KSh. 10000. The number of people that incurred a specific outpatient cost always exceeded the number of people whose hospital costs of a similar amount were covered by NHIF (figure 1).

**Figure 1.**
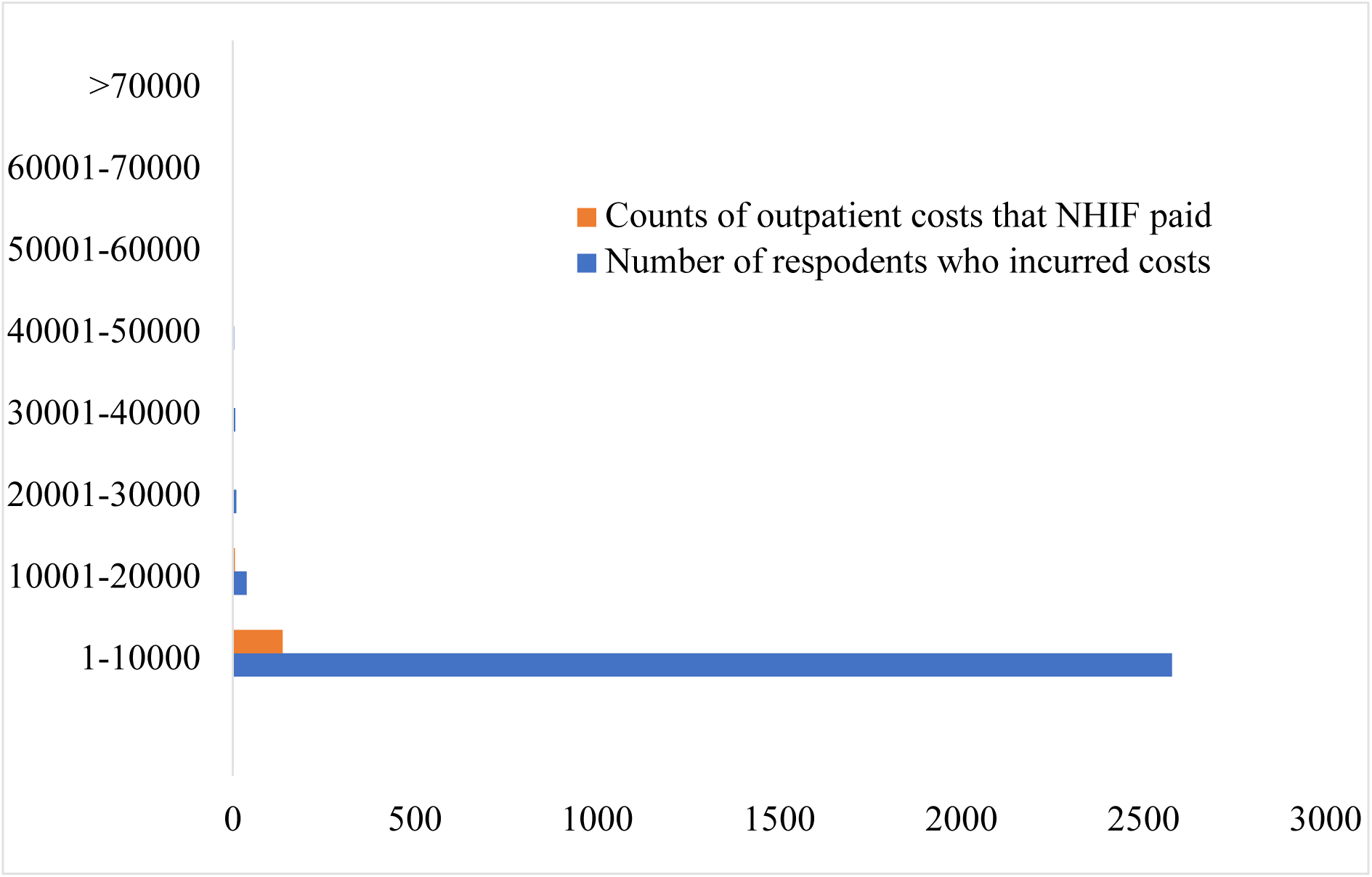
Bar chart of the counts of outpatient costs incurred versus costs that NHIF paid The total inpatient costs varied from zero Kenyan shillings (n = 62) to KSh. 4.2 million (n = 1), with 41.8% (n = 319) incurring costs exceeding KSh. 10000. Only 10.8% (n = 126) received inpatient care without paying any cash, while 29.1% paid more than KSh. 10000. Merely 162 of the inpatient bills were paid by NHIF, whereby 17.2% (n=119) were KSh. 10000 or more. Only 3.9% (n = 27) of the 701 inpatient bills were paid by private insurance, as 2.4% (n = 701) were paid in kind and five bills were paid by other means.

Individuals who incurred specific categories of costs were more than those whose similar amounts NHIF paid (Figure 2).

**Figure 2:**
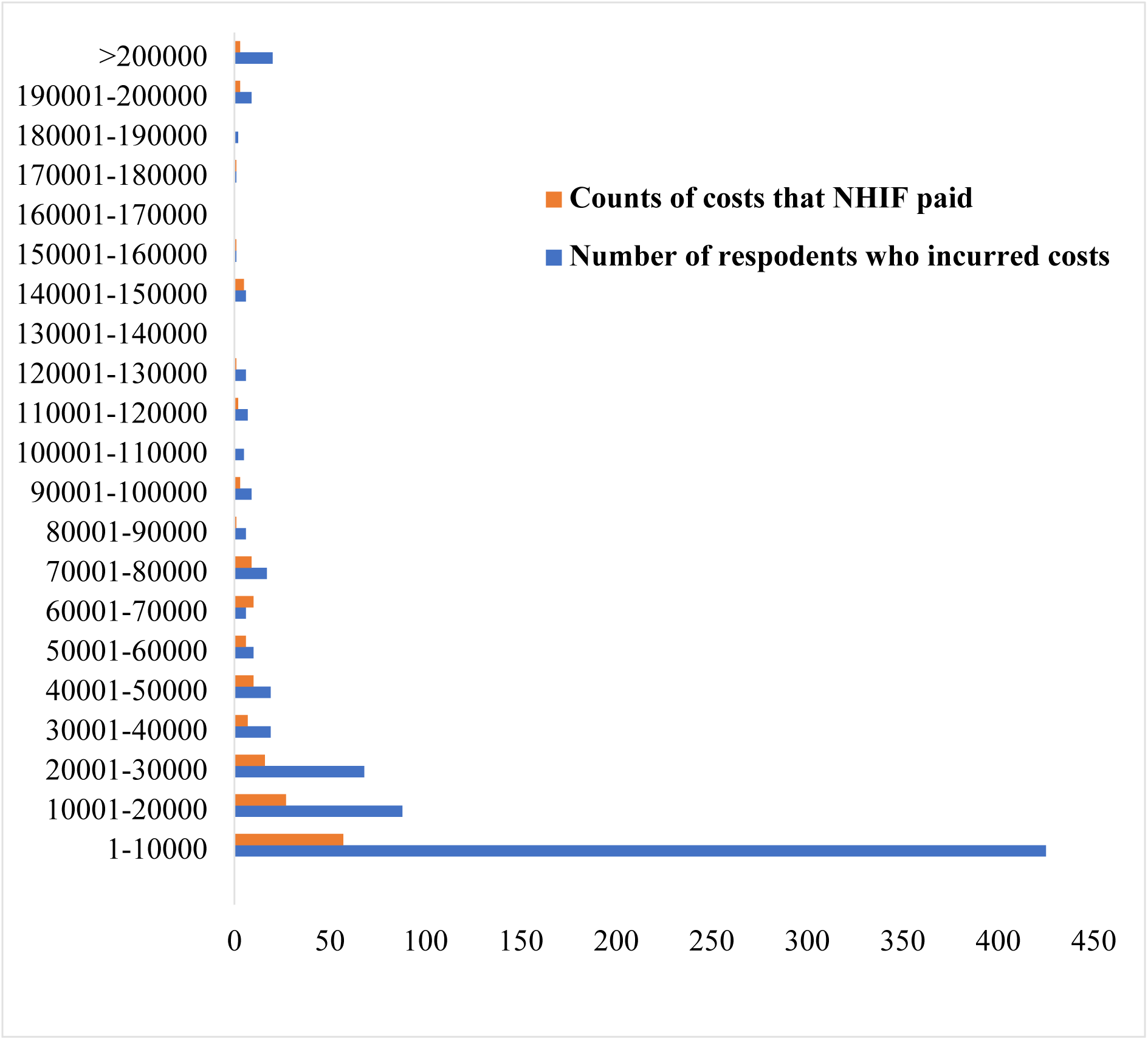
Bar graph of the counts of inpatient costs incurred versus costs that NHIF paid Both outpatient and inpatient healthcare costs in Kenya vary extensively from as low as zero to as high as KSh. 4.2 million. Most patients pay for both the outpatient and inpatient care out-of-pocket.

### Factors associated with health insurance coverage

All the factors except having paid money for the last outpatient care that were included in the bivariate analysis were statistically significantly associated with health insurance coverage (table 2).

**Table 2.** Association between characteristic of the participants and their health insurance status.

| Independent variable | Categories | Has Health Insurance % (n) | Does not have health insurance % (n) | Statistical test ($\chi^2$) | P - value |
| --- | --- | --- | --- | --- | --- |
| Gender | Male | 36.4 (1763) | 63.6 (3081) | 21.1 | 0.0001 |
|  | Female | 32.6 (3056) | 67.4 (6332) |  |  |
| Wealth | Richest | 68.8 (1870) | 31.2 (849) | 2694.4 | 0.0001 |
|  | Richer | 41.6 (1473) | 58.4 (2070) |  |  |
|  | Middle | 29.8 (788) | 70.2 (1854) |  |  |
|  | Poorer | 18.6 (424) | 81.4 (1860) |  |  |
|  | Poorest | 8.7 (264) | 91.3 (2780) |  |  |
| Educational level | Higher | 69.1 (1941) | 30.9 (870) | 2439.1 | 0.0001 |
|  | Secondary | 37.3 (1640) | 62.7 (2762) |  |  |
|  | Primary | 19.6 (1044) | 80.4 (4289) |  |  |
|  | No education | 11.5 (194) | 88.5 (1492) |  |  |
| Literacy level | Read sentence | 41.1 (4271) | 58.9 (6127) | 933.4 | 0.0001 |
|  | Read parts | 19.8 (312) | 80.2 (1264) |  |  |
|  | Cannot read | 10.5 (236) | 89.5 (2022) |  |  |
| Reads newspaper | ≥ once a week | 57.1 (825) | 42.9 (619) | 538.1 | 0.0001 |
| | $<$once a week | 43.7 (809) | 56.3 (1044) | | |
|  | Not at all | 29.1 (3185) | 70.9 (7750) |  |  |
| Has electricity | Yes | 46.7 (3653) | 53.3 (4161) | 1285.4 | 0.0001 |
|  | No | 18.2(1166) | 81.8 (5252) |  |  |
|  | No | 18.2 (1166) | 81.8 (5252) |  |  |
| Has a solar panel | Yes | 29.7 (1288) | 70.3 (3043) | 47.2 | 0.0001 |
|  | No | 35.7 (3531) | 64.3 (6370) |  |  |
| Frequency of listening to radio | ≥ once a week | 37.1 (3237) | 62.9 (5497) | 128.5 | 0.0001 |
| | $<$ once a week | 33.2 (632) | 66.8 (1274) | | |
|  | Not at all | 26.4 (950) | 73.6 (2642) |  |  |
| Frequency of watching television | ≥ once a week | 48.0 (3367) | 52.0 (3650) | 1312.5 | 0.0001 |
|  | < once a week | 28.3 (550) | 71.7 (1392) |  |  |
|  | Not at all | 17.1 (902) | 82.9 (4371) |  |  |
| Mobile phone | Yes | 37.1 (4589) | 62.9 (7771) | 448.0 | 0.0001 |
|  | No | 12.3 (230) | 87.7 (1642) |  |  |
| Smart phone | Yes | 52.1 (3240) | 47.9 (2974) | 1206.6 | 0.0001 |
|  | No | 21.9 (1349) | 78.1 (4797) |  |  |
| Has a computer | Yes | 73.8 (913) | 26.2 (324) | 965.3 | 0.0001 |
|  | No |  |  |  |  |
| Use of internet | >12 months ago | 32.3 (143) | 67.7 (300) | 1706 | 0.0001 |
|  | Last 12 months | 52.0 (3281) | 48.0 (3023) |  |  |
|  | Never | 18.6 (1395) | 81.4 (6090) |  |  |
| internet use last month | Almost daily | 58.1 (2395) | 41.9 (1729) | 1859.7 | 0.0001 |
|  | ≥ once a week | 44.0 (646) | 56.0 (822) |  |  |
|  | < once a week | 34.1 (127) | 65.9 (245) |  |  |
|  | Not at all | 20.0 (1651) | 80.0 (6617) |  |  |
| health status | Good | 34.9 (3914) | 65.1 (7315) | 40.1 | 0.0001 |
|  | Moderate | 31.4 (830) | 68.6 (1810) |  |  |
|  | Bad | 20.7 (75) | 79.3 (288) |  |  |
| Outpatient care last month | Yes | 37.7 (1251) | 62.3 (2069) | 28.8 | 0.0001 |
|  | No | 32.6(3552) | 67.4 (7329) |  |  |

### Determinants of health insurance coverage

The distribution of health insurance coverage (Yes or No) in relation to several independent variables were predicted using logistic regression. Variance inflation factors (VIF) and tolerance were examined to assess multicollinearity. The VIF values ranged from 1.0 to 4.5, hence multicollinearity was not a concern. Tolerance values ranged from 0.2 to 1.0, thus the levels of collinearity among the predictors were acceptable.

Logistic regression was conducted using the forward Wald method since there were multiple potential predictors. All the included predictors were categorical. The model converged in five iterations, indicating stability of the obtained maximum likelihood estimates. It stopped adding predictors in step 10, in which the Omnibus Test of Model Coefficients was statistically significant χ² (19) = 3736.6, *p* < .0001). From Step 0 to Step 10, the −2 Log Likelihood decreased from 15394.2 to 14483.4, indicating better fit with the added predictors. The model explained about 23.1% and 28.7% of the variance in insurance coverage status according to the Cox & Snell R² and Nagelkerke R², respectively.

In the final model (step 10), 10 variables were retained but only six were significant predictors of health insurance coverage status (table 3). Respondents with no education had 57% lower odds of health insurance coverage than individuals with higher education. The poorest respondents had 43% lower odds of insurance coverage compared to the richest participants. Similarly, individuals in households without television had 42% lower odds of health insurance compared to those whose households had television. Lacking a mobile phone or a computer were also associated with 39% and 32% lower odds of health insurance coverage, respectively. Among individuals without electricity, lacking a solar panel in the household was associated with 14% lower odds of insurance coverage.

**Table 3.** Results of the logistic regression model to identify determinants of health insurance coverage status.

| Predictor | Wald | Significance | Exp (B) | Confidence interval |
| --- | --- | --- | --- | --- |
| Education level | 206.9 | 0.0001 | 0.431 | (0.385-0.484) |
| Wealth index combined | 83.8 | 0.0001 | 0.570 | 0.505-0.643 |
| Has television | 116.7 | 0.0001 | 0.576 | 0.521-0.636 |
| Has mobile phone | 12.1 | 0.001 | 0.614 | 0.467-0.809 |
| Has a computer | 22.8 | 0.0001 | 0.683 | 0.584-0.799 |
| Household has a solar panel | 9.0 | 0.003 | 0.861 | 0.780-0.950 |

## Discussion

Only about a third of Kenyans who answered the question on insurance coverage were insured. Most Kenyans in the informal sector are uninsured, with more than 70% of the few members foregoing the renewal of their NHIF membership (Jung, 2021). Most healthcare costs were paid out-of-pocket as NHIF sometimes paid nothing or only paid a small fraction of both outpatient and inpatient bills. The NHIF’s low purchasing power despite the 2015 reforms that meant to leverage it to steer Kenya to Universal Health Coverage could be the reason for the discrepancies between the healthcare costs and NHIF payments (Mbau et al., 2020). Additionally, the premium contributions and the benefits package do not appeal to people in the informal sector, hence they choose to remain uninsured. Besides, the diversity of benefits packages with the contributors from the informal sector lacking access to pertinent services compared to civil servants demoralized Kenyans in the informal sector from contributing to NHIF (Jung, 2021).

### Low level of education predicts lack of health insurance coverage in Kenya

The likelihood of having health insurance was higher among highly educated Kenyans than Kenyans with low levels of education. The proportion of people without health insurance cover reduced with the increase in the level of education from 92% among Kenyans without education to 77% among Kenyans with tertiary education (Yego et al., 2023). The odds of health insurance coverage among individuals with higher education were nine times the odds of health insurance in a related study that analyzed the 2009 and 2014 KDHS data (Kazungu & Barasa, 2017). Active members of NHIF are more likely to be highly educated and employed while the inactive members are mostly in the informal sector (Indimuli et al., 2023). High education levels increase the chances of formal employment, and the contributions of formally employed people to NHIF are compulsory. Therefore, the attainment of universal health coverage in Kenya through premium-based health insurance is dependent on Kenya’s progress in attaining high education levels and creating formal jobs.

### Poverty predicts lack of health insurance in Kenya

Health insurance uptake corresponds to the relative wealth levels of Kenyans, with individuals in the lowest wealth levels having a low likelihood of NHIF enrolment (Wanjiku, 2024). Unaffordability of premiums by Kenyans in the informal sector could be contributing to financing constraints by NHIF (Mbau et al., 2020), hence the NHIF’s inability to pay bills matching the healthcare costs that Kenyans incur. Only a small fraction of Kenyans in the informal sector could afford the Sh. 500 monthly premium, which was a sharp increase from the initial Sh. 160, yet the cost of living continually increases as low incomes persist in the informal sector (Jung, 2021). High-earning people in the informal sectors, who also mostly belong to social associations such as Matatu Savings and Credit Cooperatives (SACCOs), are more likely to be active members of NHIF compared to wage workers and self-employed small business operators (Indimuli et al., 2023). The ability to pay the premiums of an health insurance scheme determines whether people will contribute to it (Jung, 2021). Relying on payment of premiums to achieve universal health coverage may be a tall order in Kenya due to the large number of Kenyans living below the poverty line (Kazungu & Barasa, 2017).

Therefore, creating a supportive business environment that will increase earnings in the informal sector and enhancing social welfare programs for the vulnerable individuals are necessary to increase the health insurance coverage if Kenya will rely on the premium-based health insurance to achieve universal health coverage (Wanjiku, 2024).

### Lacking a television predicts lack of health insurance coverage in Kenya

Media factors such as watching television influence the demand for health insurance in developing countries (Musoke et al., 2022). NHIF mainly used television and radio to communicate the revised premiums and benefits package to the public (Mbau et al., 2020).

However, the revised benefits package after 2015 NHIF reforms were seemingly inadequately communicated to the public (Mbau et al., 2020), hence non-registered Kenyans in the informal sector may not have been convinced to be NHIF members. Although 97% of Kenyans know of NHIF, several people in the informal sector lack adequate levels of knowledge about health insurance schemes, hence they do not enlist or contribute to NHIF (Indimuli et al., 2023). People in the informal sector require better awareness of the services they would access if they were contributing to the social health insurance fund for evidence-based decision-making of their uptake of the insurance cover (Yego et al., 2023). Therefore, more awareness initiatives through audio-visual channels such as the television and social media are crucial in the uptake of health insurance.

### Lacking a phone or a computer predicts lack of health insurance coverage in Kenya

Digital access tools influence the uptake of health insurance in Kenya. A similar study reported a high likelihood of lack of health insurance coverage among more Kenyans without mobile phones compared to Kenyans with mobile phones. NHIF partly communicated its reforms through social media, website, and short message service (SMS), which require ownership of a mobile phone to access (Mbau et al., 2020). Hence, the low likelihood of insurance coverage among Kenyans without phones and computers could be due to limited access to health insurance-related information. The digital customer interaction strategy at NHIF relied on the ownership of mobile phones or computers by Kenyans, and it influenced service delivery to beneficiaries as the public sought information on the benefit packages and premium payments (Wachira & Gichuhi, 2023). Besides, considering the high rate of using MPesa to make payments including premium remittances to NHIF (Kirika, 2018), lack of a mobile phone may have impeded active membership to NHIF.

The main limitation of this study is that it used a cross-sectional design study design, hence causality cannot be inferred. However, influence of the predictors on the health insurance coverage status can be reliably deduced by contextualizing the findings to related literature. Secondly, the researchers were not in control of data collection since it was pre-collected in KDHS. Nevertheless, the KDHS was the only feasible way to obtain quality data from a nationally-representative study.

## Conclusion

Low uptake of NHIF coverage in Kenya persisted in 2022 based on the 2022 KDHS survey; it may be among the impetus behind the government of Kenya’s transition to SHA. The main predictors of low levels of NHIF coverage based on the KDHS survey include low level of education, poverty, lack of a television, and lack of a phone or a computer. SHA should amplify the interventions that worked in NHIF such as awareness campaigns on television, radio, and social media since most Kenyans have televisions and mobile phones. It should consider establishing a communication strategy to inform uneducated people and those who rarely listen to radio, watch television or own mobile phones or computers to ensure every Kenyan knows about the SHA requirements and benefits. It should establish precise and reliable mechanisms of identifying all poor Kenyans to exempt them from payment of SHIF premiums. Possibly, the government of Kenya should consider adopting a fully tax-funded social health insurance scheme considering the large number of Kenyans with low wealth levels, which complicates collection of substantial premiums. Improving education levels and economic performance coupled with practically enriching the health insurance package can increase uptake of health insurance even if the premium-based model is retained.

## Data Availability

The data analyzed in this study is deposited in Mendeley Data, a publicly accessible data repository. The DOI for the dataset will be made available as soon as Mendeley Data generates it.

## Acknowledgments

We acknowledge the Kenya National Data Archive (KeNADA) for granting access to the Kenya Demographic and Health Survey 2022 datasets.

## Author contributions

Conceptualization, E.A., M.A., and D.K.; Methodology, E.A., M.A., and D.K.; Validation, E.A., M.A., and D.K.; Formal Analysis, D.K.; Investigation, D.K.; Resources, E.A. and M.A.; Data Curation, E.A., M.A., and D.K.; Writing – Original Draft Preparation, D.K.; Writing – Review & Editing, E.A., M.A., and D.K.; Visualization, D.K.; Supervision, D.K.; Project Administration, E.A.

## Ethical considerations

This study was conducted using publicly available data, hence it did not require ethical review.

## Funding

The study was self-funded by the authors.

## Competing interests

The authors declare no competing interests

## Notes

### Competing Interest Statement

The authors have declared no competing interest.

### Funding Statement

The author(s) received no specific funding for this work.

